# Parental COVID-19 vaccine hesitancy and vaccine uptake among children and adolescents in the US: Findings from a prospective national cohort

**DOI:** 10.1101/2022.10.16.22281135

**Authors:** Madhura S. Rane, McKaylee Robertson, Drew Westmoreland, Rebecca Zimba, Sarah G. Kulkarni, Yanhan Shen, Amanda Berry, Mindy Chang, William You, Christian Grov, Denis Nash, the CHASING COVID Cohort Team

## Abstract

**Objectives:** Our aim was to measure COVID-19 vaccine uptake among children aged 5-17 years old via parents participating in the CHASING COVID Cohort and identify sociodemographic factors associated with it.

**Methods:** In this longitudinal study, parents of school-aged children were asked about their own vaccination status and that of their children at three time points between June 2021-January 2022, along with reasons for vaccinating immediately or delaying vaccinations for their children. Multivariable log binomial models were used to identify correlates of vaccine uptake among children.

**Results:** Of the 1,583 children aged 5-17 years, 64.9% were vaccinated. Over 40% of parents of 5-11 year old children who intended to delay vaccinating their child in June 2021 had still not vaccinated them by January 2022, including 30% of the parents who were vaccinated. After adjusting for measured confounders, parents’ vaccination status was associated with higher likelihood of children’s vaccine uptake (age-specific adjusted odds ratios [aORs]: aOR_16-17_ 3.7, 95% CI 2.3, 5.9, aOR_12-15_ 3.7, 95% CI 2.6, 5.3; aOR_5-11_ 10.6, 95% CI 5.4, 20.9). Parents’ education (aOR_16-17_ 1.4, 95% CI 1.1, 1.8, aOR_12-15_ 1.5, 95% CI 1.2, 1.9; aOR_5-11_ 2.1, 95% CI 1.5, 2.9) and worry about others getting infected (aOR_5-11_ 1.4, 95% CI 1.1, 1.6) were also associated with higher vaccine uptake among children. A higher proportion of parents of 5-11 year olds (vs. 12-17 year olds) had concerns about vaccine safety and effectiveness.

**Conclusion:** To increase vaccination coverage among young children, vaccination campaigns should focus on both vaccinated and unvaccinated parents and messaging should be specific to the child’s age.

## Introduction

On May 10th, 2021, the Centers for Disease Control and Prevention (CDC) Advisory Committee of Immunization Practices (ACIP) recommended the Pfizer-BioNTech COVID-19 vaccine for 12-15 year olds under Emergency Use Authorization (EUA).^1^ This was soon followed by EUA approval for 5-11 year olds on November 2nd, 2021.^2^ However, pediatric vaccine uptake in the United States (US) has been low, especially in the younger age groups. In February 2022, only 27% of 5-11 year olds were estimated to have been fully vaccinated against COVID-19 compared to 64% of 12-17 year olds.^3^ In January 2022, the weekly incidence of COVID-19 was higher among children <17 years compared to adults >50 years, and hospitalization rates among 0-17 year olds were the highest they have been since the beginning of the pandemic.^4^ Vaccination remains the most effective strategy to protect against severe disease and increasing COVID-19 vaccination coverage among children might also indirectly protect vulnerable older adults by reducing community transmission, as has been shown in influenza vaccination studies.^5,6^

Using longitudinal data from the national CHASING COVID Cohort, we measured trends in vaccine uptake among participants’ children aged 5-17 years old. We assessed the association between parental vaccination status and vaccine uptake among children and identified other sociodemographic, child-related, and COVID-19 related risk factors associated with pediatric vaccine uptake.

## Methods

The Communities, Households, and SARS-CoV-2 Epidemiology COVID (CHASING COVID) Cohort study^7^ is a national prospective cohort study launched in March, 2020 among adults ≥18 years residing in the U.S. to understand the spread and impact of the SARS-CoV-2 pandemic within households and communities. As of January 2022, 9 full interview rounds were completed, which captured longitudinal information on participant demographics, COVID-related exposures, outcomes, detailed symptoms, non-pharmaceutical intervention (NPI) use, and vaccine uptake. Questionnaires are available on the study’s homepage (https://cunyisph.org/chasing-covid/). The study was approved by the Institutional Review Board at the City University of New York (CUNY).

For this study, we included participants who were parents of children aged 5-17 years and who responded to at least one of three survey rounds conducted in June 2021, October 2021, and January 2022. At each time point, parents were asked whether vaccine-eligible child/children (grouped as 16-17 years, 12-15 years, and 5-11 years old) in their household had received at least 1 dose of an FDA-approved COVID-19 vaccine. Child’s vaccination status (vaccinated/unvaccinated) as of January 2022 was determined based on the latest survey completed. Parents with children in different vaccine-eligible age groups responded separately for each child. However, parents could only provide one response if they had multiple children in the same age group. Given EUA timing, parents were only asked about the vaccination status of 5-11 year old children in the January 2022 survey round. In each survey, parents of children who were not yet vaccinated (including those ineligible for the vaccine at the time of the survey) were asked about their willingness to vaccinate them (yes/no). Parents were asked about their reasons for vaccinating or not vaccinating their children.

The primary outcome was vaccination status of children, defined as *receiving at least one dose of any COVID-19 vaccine*, in age categories 5-11 years, 12-15 years and 16-17 years as of January 2022. The main exposure was parental vaccination status determined using longitudinally collected vaccination data for parents and dichotomized as vaccinated vs. unvaccinated at the time of reporting of child vaccination status. Other correlates of interest, such as, having a younger sibling, in-person vs remote/hybrid school attendance, whether parents were worried about their loved ones getting infected, prior SARS-CoV-2 in infection in parents, knowing someone who died of COVID, parents’ employment status, and income loss due to having to provide childcare, were collected in each survey round and were analysed as time updated covariates. Parents’ age, gender, race/ethnicity, and education attainment were collected at baseline.

Sankey plots were used to visualize the trend in parental willingness for their children to be vaccinated. Univariate log-binomial regression models were used to identify parent- and child-related factors independently associated with vaccine uptake among children, with statistical significance defined as 2-sided P <0.05. Multivariable log-binomial models were fit to assess the association between parental vaccination status and child’s vaccine uptake for each age group separately, adjusted for parent age, gender, race/ethnicity, education, and employment. Separate multivariable models were also fit for associations of parental education attainment, worry about COVID, knowing someone who died from COVID, income loss due to childcare, and child’s school attendance type with child’s vaccine uptake, each adjusted for confounders selected a priori. All analyses were conducted in R Studio (version 4.0.2).

## Results

The analyses included 1,208 parents (mean [SD] age, 39.1 [9.8] years) with 1,583 children aged 5 to 17 years in the household. As of January 2022, 252 (72.2%) children aged 16-17 years, 406 (69.8%) children aged 12-15 years, and 370 (56.6%) children aged 5-11 years had received at least 1 dose of their vaccine (Figure 1). Roughly 64% of parents of 16-17 year olds, 74% of parents of 12-15 year olds and 61% of parents of 5-11 year olds who responded that they would immediately vaccinate their child in June 2021 had vaccinated their child by January 2022 (Figure 1). Of those who said they would delay vaccinations for their children in June 2021, 45% of parents of 16-17 year olds, 44% of parents of 12-15 year olds, and 67% of parents of 5-11 year olds had still not vaccinated them by January 2022.

**Figure 1:**
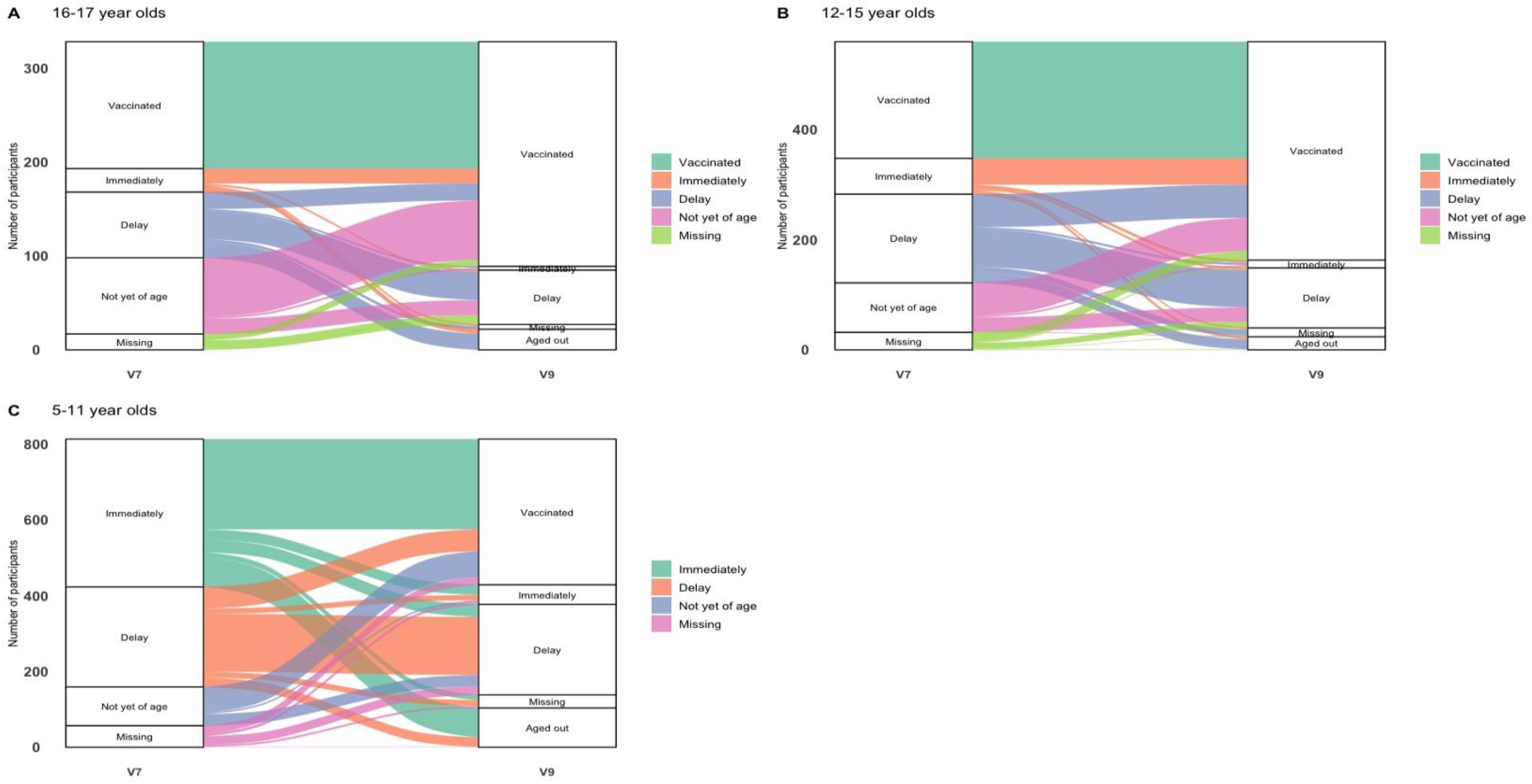
Sankey plot showing trajectory of parents intention to vaccinate children in June 2021 to vaccination status of children in January 2022 among parents of 5-17 year old children in the CHASING COVID cohort.

Of the 1,583 children whose vaccination status was reported by their parents, 35% (N=555) had not received any COVID-19 vaccine dose as of January 2022. Among vaccinated parents, the proportion who had not vaccinated their children by January 2022 was 16.8% (N = 129) and 29.7% (N =153) for children aged 12-17 and 5-11 years old, respectively (Table 1). For 12-17 year olds, only 2% (17/735) of unvaccinated parents reported that their children were vaccinated, while for 5-11 year olds, this proportion was 6% (8/138).

**Table 1:** Characteristics of parents by age group and vaccination status of children.

|  | 5-11 years old |  | 12-15 years old |  | 16-17 years old |  |
| --- | --- | --- | --- | --- | --- | --- |
|  | Vaccinated<br>(n= 370), N(%) | Unvaccinated<br>(n=283), N(%) | Vaccinated<br>(n=406), N(%) | Unvaccinated<br>(n=175), N(%) | Vaccinated<br>(n=252), N(%) | Unvaccinated<br>(n=97), N(%) |
| **Characteristics** |  |  |  |  |  |  |
| **Parent vaccination status** |  |  |  |  |  |  |
| **Vaccinated** | 362 (97.84) | 153 (54.06) | 393 (96.80) | 82 (46.86) | 243 (96.43) | 47 (48.45) |
| **Unvaccinated** | 8 (2.16) | 130 (45.94) | 13 (3.20) | 93 (53.14) | 9 (3.57) | 50 (51.55) |
| **Age group** |  |  |  |  |  |  |
| **30-39** | 180 (48.65) | 163 (57.60) | 143 (35.22) | 92 (52.57) | 67 (26.59) | 38 (39.18) |
| **18-29** | 52 (14.05) | 63 (22.26) | 38 (9.36) | 20 (11.43) | 20 (7.94) | 9 (9.28) |
| **40-49** | 117 (31.62) | 41 (14.49) | 161 (39.66) | 44 (25.14) | 104 (41.27) | 33 (34.02) |
| **50-59** | 11 (2.97) | 8 (2.83) | 54 (13.30) | 13 (7.43) | 51 (20.24) | 12 (12.37) |
| **60+** | 10 (2.70) | 8 (2.83) | 10 (2.46) | 6 (3.43) | 10 (3.97) | 5 (5.15) |
| **Gender** |  |  |  |  |  |  |
| **Male** | 145 (39.19) | 71 (25.09) | 152 (37.44) | 57 (32.57) | 76 (30.16) | 28 (28.87) |
| **Female** | 219 (59.19) | 210 (74.20) | 247 (60.84) | 118 (67.43) | 175 (69.44) | 69 (71.13) |
| **Non-binary** | 6 (1.62) | 2 (0.71) | 7 (1.72) | 0 (0.00) | 1 (0.40) | 0 (0.00) |
| **Race/Ethnicity** |  |  |  |  |  |  |
| **White** | 186 (50.27) | 126 (44.52) | 216 (53.20) | 83 (47.43) | 133 (52.78) | 44 (45.36) |
| **Other races\*** | 182 (49.19) | 157 (55.48) | 189 (46.55) | 92 (52.57) | 118 (46.83) | 53 (54.64) |
| **Education** |  |  |  |  |  |  |
| **Less than high school** | 29 (7.84) | 77 (27.21) | 348 (85.71) | 110 (62.86) | 35 (13.89) | 34 (35.05) |
| **High school or higher** | 341 (92.16) | 206 (72.79) | 58 (14.29) | 65 (37.14) | 217 (86.11) | 63 (64.95) |
| **Employment** |  |  |  |  |  |  |
| **Out of work** | 21 (5.68) | 23 (8.13) | 32 (7.88) | 21 (12.00) | 28 (11.11) | 16 (16.49) |
| **Employed** | 301 (81.35) | 195 (68.90) | 314 (77.34) | 117 (66.86) | 189 (75.00) | 60 (61.86) |
| **Homemaker** | 31 (8.38) | 56 (19.79) | 46 (11.33) | 27 (15.43) | 29 (11.51) | 14 (14.43) |
| **Student** | 7 (1.89) | 5 (1.77) | 3 (0.74) | 5 (2.86) | 2 (0.79) | 3 (3.09) |
| **Retired** | 9 (2.43) | 3 (1.06) | 11 (2.71) | 5 (2.86) | 4 (1.59) | 4 (4.12) |
| **Younger sibling in Household** |  |  |  |  |  |  |
| **Yes** | 215 (58.11) | 187 (66.08) | 274 (67.49) | 126 (72.00) | 147 (58.33) | 57 (58.76) |
| **No** | 155 (41.89) | 96 (33.92) | 132 (32.51) | 49 (28.00) | 105 (41.67) | 40 (41.24) |
| **School attendance status of children** |  |  |  |  |  |  |
| **All school age children attend in person** | 236 (63.78) | 188 (66.43) | 158 (38.92) | 92 (52.57) | 103 (40.87) | 48 (49.48) |
| **All school age children attend remotely** | 32 (8.65) | 20 (7.07) | 82 (20.20) | 21 (12.00) | 50 (19.84) | 12 (12.37) |
| **All children homeschooled** | 14 (3.78) | 15 (5.30) | 19 (4.68) | 11 (6.29) | 8 (3.17) | 8 (8.25) |
| **Attend a hybrid model** | 39 (10.54) | 31 (10.95) | 96 (23.65) | 20 (11.43) | 60 (23.81) | 8 (8.25) |
| **Other/NA** | 49 (13.24) | 29 (10.25) | 51 (12.56) | 31 (17.71) | 31 (12.30) | 21 (21.65) |
| **Worry about others getting infected** |  |  |  |  |  |  |
| **Yes** | 291 (78.65) | 184 (65.02) | 258 (63.55) | 102 (58.29) | 158 (62.70) | 61 (62.89) |
| **No** | 78 (21.08) | 98 (34.63) | 148 (36.45) | 73 (41.71) | 94 (37.30) | 36 (37.11) |
| **Parent previously infected** |  |  |  |  |  |  |
| **Yes** | 92 (24.86) | 65 (22.97) | 87 (21.43) | 45 (25.71) | 60 (23.81) | 18 (18.56) |
| **No** | 278 (75.14) | 218 (77.03) | 319 (78.57) | 130 (74.29) | 192 (76.19) | 79 (81.44) |
| **Parents know** |  |  |  |  |  |  |
| **someone who died**<br>**of COVID** |  |  |  |  |  |  |
| **Yes** | 206 (55.68) | 130 (45.94) | 245 (60.34) | 88 (50.29) | 145 (57.54) | 56 (57.73) |
| **No** | 164 (44.32) | 153 (54.06) | 161 (39.66) | 87 (49.71) | 107 (42.46) | 41 (42.27) |
| **Income loss due to**<br>**childcare (ref:No)** |  |  |  |  |  |  |
| **Yes** | 66 (17.84) | 74 (26.15) | 73 (17.98) | 36 (20.57) | 145 (57.54) | 56 (57.73) |
| **No** | 304 (82.16) | 209 (73.85) | 333 (82.02) | 139 (79.43) | 107 (42.46) | 41 (42.27) |

\*Other races include participants who identified as non-Hispanic Black, Hispanic, Asian, Pacific
Islander, and other

**Table 2:** Multivariable regression models for factors associated with vaccination uptake in children.

|  | 5-11 year olds<br>(n=653) |  | 12-15 year olds<br>(n=581) |  | 16-17 year olds<br>(n=349) |  |
| --- | --- | --- | --- | --- | --- | --- |
| Characteristics | aOR | 95% CI | aOR | 95% CI | aOR* | 95% CI |
| **Parent vaccination status\*** | **10.61** | **5.38, 20.89** | **3.73** | **2.63, 5.30** | **3.67** | **2.29, 5.86** |
| **Education\*\*** |  |  |  |  |  |  |
| Less than high school | ref | ref | ref | ref | ref | ref |
| High school or higher | **2.08** | **1.51, 2.86** | **1.53** | **1.25, 1.87** | **1.41** | **1.09, 1.81** |
| **School attendance status of children\*\*\*** |  |  |  |  |  |  |
| All school age children attend in person | ref | ref | ref | ref | ref | ref |
| All school age children attend remotely | 1.12 | 0.98, 1.28 | **1.11** | **1.03, 1.19** | 1.03 | 0.92, 1.17 |
| All children homeschooled | 0.97 | 0.70, 1.34 | 0.97 | 0.77, 1.23 | 0.81 | 0.52, 1.23 |
| Attend a hybrid model | 0.88 | 0.72, 1.08 | **1.1** | **1.02, 1.19** | **1.12** | **1.03, 1.21** |
| **Worry about others** | **1.36** | **1.15, 1.60** | 1.05 | 0.96, 1.15 | 1.08 | 0.97, 1.18 |
| **getting infected\*\*\*\***<br>**(ref=No)** |  |  |  |  |  |  |
| **Parents know someone**<br>**who died of COVID\***<br>**(ref:No)** | **1.14** | **1.01, 1.28** | 1.1 | 1.00, 1.21 | 0.98 | 0.87, 1.10 |
| **Income loss due to**<br>**childcare\* (ref:No)** | 0.86 | 0.73, 1.02 | 1.02 | 0.89, 1.16 | 0.99 | 0.79, 1.22 |

\*Adjusted for parent age, gender, race/ethnicity, education, employment
\*\*Adjusted for parent age, gender, race/ethnicity, employment
\*\*\*Adjusted for parent age, gender, race/ethnicity, education, employment, parent vaccination status, parent worry about others getting COVID-19
\*\*\*\*Adjusted for parent age, gender, race/ethnicity, education, employment, knowing someone who died of COVID-19, parent previously infected with COVID-19

In univariate models, parental vaccination uptake, higher parental age, and higher education attainment was associated with a higher likelihood of vaccine uptake for children in all age groups (Supplementary table 1). Female parents of 5-11 year old children were less likely to report that their child was vaccinated. Having a sibling younger than 5 years in the household and loss of income for parents due to unavailability of childcare was also associated with lower vaccine uptake in 5-11 year olds. Parents who reported being worried about others getting infected with SARS-CoV-2 and knowing someone who died of COVID were more likely to report that their 5-11 year olds were vaccinated. Type of school attendance (fully in-person, hybrid, fully remote) was independently associated with adolescent vaccine uptake in univariate models (Supplementary table 1).

In multivariable models, the odds of parental vaccination uptake was highly correlated with child’s vaccine uptake in all child age groups, after adjusting for parent age, gender, race/ethnicity, education, and employment (aOR_16-17_ 3.67, 95% CI 2.29, 5.86, aOR_12-15_ 3.73, 95% CI 2.63, 5.30; aOR_5-11_ 10.61, 95% CI 5.38, 20.89). Parents with education of high school or higher were more likely than their counterparts to report that their children were vaccinated in all child age groups (aOR_16-17_ 1.41, 95% CI 1.09, 1.81, aOR_12-15_ 1.53, 95% CI 1.25, 1.87; aOR_5-11_ 2.08, 95% CI 1.51, 2.86). Adolescents who attended schools following a hybrid model of instruction compared to those who attended school fully in-person were slightly more likely to be vaccinated (aOR_16-17_ 1.12, 95% CI 1.03, 1.21, aOR_12-15_ 1.10, 95% CI 1.02, 1.19). Worry among parents about others contracting COVID-19 (aOR 1.36, 95% CI 1.15, 1.60) and knowing someone who died of COVID-19 (aOR 1.14, 95% CI 1.01, 1.28) was associated with higher vaccine uptake for younger children aged 5-11, but not for older children.

### Parental reasons for vaccinating or not vaccinating children

Across age groups, parents whose children were vaccinated reported that protecting their children against COVID-19, concern about variants such as Delta and Omicron and belief that the vaccine is effective motivated them to get their children vaccinated (Figure 2). On the other hand, parents whose children were not vaccinated reported concerns regarding the long and short term effects of the vaccine, as well as about vaccine effectiveness. Among those whose children were vaccinated, compared to parents of 5–11-year-olds, parents of 12–17-year-olds were more likely to report vaccinating their children because their child wanted the vaccine (45% vs. 55%). Compared to parents of 5–11-year-olds, parents of 12–17-year-olds were less likely to report concerns about new variants (77% vs. 65%) or a pediatrician’s recommendation (36% vs. 28%) as reasons for vaccinating children. Among those whose children were not vaccinated, parents of 5–11-year-olds were more likely to report lack of full FDA approval (18% vs. 5%), long-term (49% vs. 31%) and short-term vaccine (36% vs. 19%) side effects, and concerns about vaccine effectiveness (33% vs. 20%) as reasons for not getting their children vaccinated (Figure 2).

**Figure 2:**
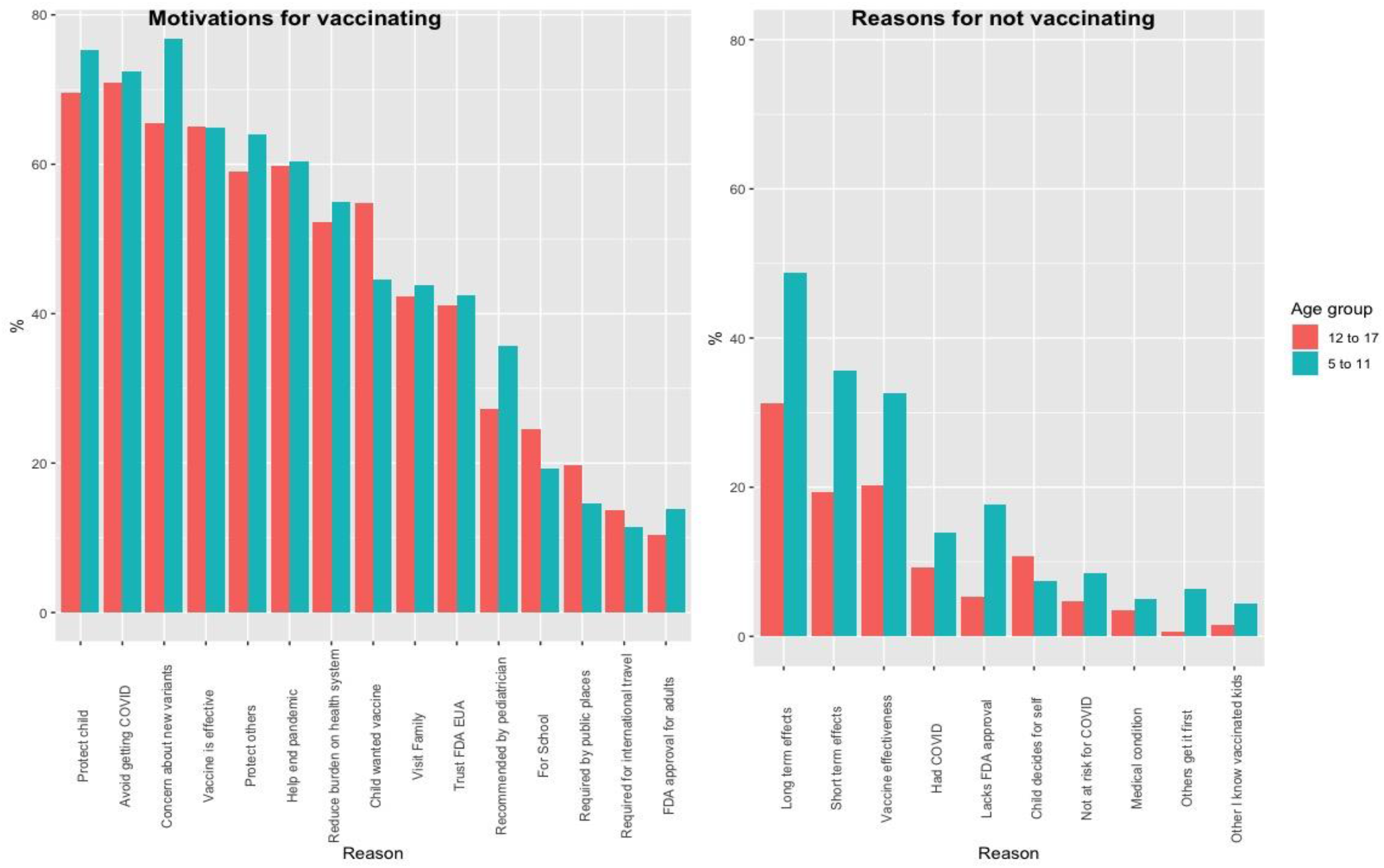
Motivation for vaccinating and reasons for not vaccinating children among parents of 5-17 year olds in the CHASING COVID cohort, January 2022

## Discussion

We prospectively examined COVID-19 vaccine uptake among children aged 5-17 years and the role of parent vaccination status as a factor that may strongly influence vaccine uptake comparing young children to adolescents in a U.S. national cohort. COVID-19 vaccine uptake was lower among 5-11 year olds compared to 12-17 year olds as of January 2022. Parental vaccination status was strongly and positively associated with vaccination uptake in children of all age groups; however, even among vaccinated parents of 5-11 year olds, one-third had not yet vaccinated their vaccine-eligible children. A substantial proportion (69%) of parents who expressed hesitancy to vaccinate their children in previous survey rounds still had not vaccinated their children by January 2022. Parents of younger children were most concerned about vaccine-related side-effects and vaccine effectiveness compared to parents of adolescents. Improving pediatric vaccination rates in the U.S. will require addressing drivers of vaccine hesitancy among both vaccinated and unvaccinated parents with attention to messaging that is specific to the child’s age. Full FDA approval would also likely help to increase uptake among children in the youngest age group.

In an earlier report from this cohort that examined parental hesitancy for pediatric vaccination, we found that parents being vaccinated was associated with a greater intention to vaccinate children, when they become eligible.^8^ In this prospective study, and similar to other cross-sectional studies in the U.S.^9,10^, parents’ vaccination status was the strongest predictor of vaccine uptake in their children, and this result was consistent across age groups of children. Still, as high as 29% of parents who were vaccinated had chosen to not vaccinate their 5-11 year old children as of January 2022. Given that most U.S. adults are vaccinated, the absolute number of unvaccinated children of vaccinated parents is likely to be quite high. Thus, COVID-19 pediatric vaccination awareness should focus on parents of younger children regardless of their vaccination status to boost overall pediatric vaccine uptake.

A substantial proportion of parents who were unwilling to vaccinate their children in previous rounds remained hesitant, especially if their children were young.^8^ This trend was observed even among parents who were themselves vaccinated. We found that concerns about safety and effectiveness of COVID-19 vaccines were heightened for parents of 5-11 year olds compared to parents of older children. Tailoring messaging about safety of COVID-19 vaccines to parents of young children and educating them about the risks of COVID-19 infection to young children is crucial to improve vaccine uptake in this age group. This is particularly important now that COVID-19 vaccines are recommended for children 6 months - 4 years as concerns about vaccine safety are likely to be the highest for this group.

We found that parents of children 12-17 years old, that is children who attended middle or high school, were more likely to get their children vaccinated if they attended school remotely or partly remotely. Now that school-wide mask mandates are being lifted^11^, vaccinating school-going children is of prime importance, as they may help to reduce transmission events in congregated settings like classrooms, which could lead to interruptions in learning and other social activities. ^12^

Studies have also shown that high COVID-19 risk perception and previous experience with COVID-19 such as previous infection or loss of a family member due to COVID are associated with lower vaccine hesitancy among adults ^13–15^. In keeping with these findings, in our study too, parents of 5-11 year olds who reported knowing someone who died of COVID-19 or worried about others becoming infected with COVID were more likely to have their children vaccinated. Parents with previous COVID-19 infection were more likely to vaccinate younger children, but this association was marginally significant. These results suggest that direct personal experience with COVID-19 might influence parents’ decision to either vaccinate their children immediately or delay vaccination.

Our study has some limitations. Responses were self-reported, introducing the possibility of recall bias. While our participants are geographically and sociodemographically diverse, they are research volunteers and thus not representative of the general population, potentially limiting generalizability. Because recruitment was done online, parents without internet access were not able to participate in the cohort. Although we adjusted for socidemographic factors in all models, there is a possibility of unmeasured confounding, which could explain some of our observed associations. Finally, older children became eligible for COVID-19 vaccination well before younger children, which could play a role in lower vaccine uptake among 5-11 year olds during our study timeframe.

Low vaccine uptake among children puts them at a high risk of severe disease and with the possibility of development of long COVID ^16,17^, both outcomes that are highly preventable with vaccination. Higher vaccination uptake could also have implications for reducing overall transmission in the community, which helps protect vulnerable people in households and communities of children for whom vaccine-derived immunity may fall short. Transparent and evidence informed-messaging by public health agencies about safety of COVID-19 vaccines among children, particularly to vaccinated and unvaccinated parents of younger children, is critical to increase vaccine uptake and address parents’ concerns about vaccine adverse effects. Vaccine manufacturers and the FDA should prioritize and accelerate their process of providing full FDA approval to pediatric vaccines to improve vaccine confidence among parents. Without deliberate action, vaccine uptake among children under 5 is likely to be even lower. Future research should focus on changes in vaccination uptake when COVID-19 vaccines receive full approval.

## Data Availability

All data produced in the present study are available upon reasonable request to the authors.

https://zenodo.org/record/6127735#.Y0v2Ny-B1H_

