## Supplementary Tables for "Parental COVID-19 vaccine hesitancy and vaccine uptake among children and adolescents in the US: Findings from a prospective national cohort"

sTable 1: Univariate regression models for factors associated with vaccination uptake in children

| | **16-17 year olds (n=358)** | | **12-15 year olds (n=599)** | | **5-11 year olds (n=679)** | |
|---|---|---|---|---|---|---|
| **Parental characteristics** | OR | 95% CI | | | OR | 95% CI |
| **Parent vaccinated** | **3.81** | **2.52, 6.45** | **3.88** | **2.81, 5.69** | **12.12** | **6.66, 26.15** |
| **Age group** | | | | | | |
| 30-39 | ref | ref | ref | ref | ref | ref |
| 18-29 | 1.08 | 0.77, 1.39 | 1.07 | 0.85, 1.31 | 0.86 | 0.67, 1.06 |
| 40-49 | **1.18** | **1.00, 1.42** | **1.29** | **1.14, 1.46** | **1.41** | **1.22, 1.61** |
| 50-59 | **1.26** | **1.04, 1,53** | **1.32** | **1.11, 1.53** | 1.1 | 0.67, 1.52 |
| 60+ | 1.04 | 0.64, 1.43 | 1.02 | 0.62, 1.39 | 1.05 | 0.62, 1.49 |
| **Gender** | | | | | | |
| Male | ref | ref | ref | ref | ref | ref |
| Female | 0.98 | 0.85, 1.14 | 0.93 | 0.83, 1.04 | **0.76** | **0.66, 0.86** |
| Non-binary | NA | NA | NA | NA | 1.11 | 0.60, 1.45 |
| **Race/Ethnicity** | | | | | | |
| White | ref | ref | ref | ref | ref | ref |
| non-White | 0.91 | 0.80, 1.04 | 0.93 | 0.83, 1.04 | 0.9 | 0.78, 1.03 |
| **Education** | | | | | | |

| | | | | | | |
|---|---|---|---|---|---|---|
| Less than high school | ref | ref | ref | ref | ref | ref |
| High school or higher | **1.52** | **1.22, 1.99** | **1.61** | **1.34, 1.98** | **2.27** | **1.70, 3.21** |
| **Employment** | | | | | | |
| Out of work | ref | ref | ref | ref | ref | ref |
| Employed | 1.19 | 0.97, 1.56 | 1.2 | 0.98, 1.56 | 1.27 | 0.96, 1.82 |
| Homemaker | 1.05 | 0.77, 1.45 | 1.04 | 0.79, 1.40 | 0.74 | 0.49, 1.15 |
| Student | 0.62 | 0.12, 1.34 | 0.62 | 0.17, 1.24 | 1.22 | 0.61, 2.04 |
| Retired | 0.78 | 0.29, 1.37 | 1.13 | 0.70, 1.63 | 1.57 | 0.92, 2.43 |
| **Younger sibling in HH (ref:No)** | 0.99 | 0.87, 1.14 | 0.93 | 0.84, 1.05 | **0.86** | **0.75, 0.99** |
| **School attendance status of children** | | | | | | |
| All school age children attend in person | ref | ref | ref | ref | ref | ref |
| All school age children attend remotely | 1.94 | 0.97, 4.12 | **1.25** | **1.09, 1.43** | 1.1 | 0.85, 1.25 |
| All children homeschooled | 0.46 | 0.16, 1.33 | 1 | 0.71, 1.28 | 0.86 | 0.55, 1.20 |
| Attend a hybrid model | **3.49** | **1.62, 8.42** | **1.3** | **1.15, 1.48** | 1 | 0.78, 1.22 |

| | | | | | | |
|---|---|---|---|---|---|---|
| **Worry about others getting infected (ref=No)** | 0.99 | 0.87, 1.14 | 1.07 | 0.95, 1.20 | **1.38** | **1.16, 1.67** |
| **Parent previously infected (ref:No)** | 1.08 | 0.92, 1.24 | 0.92 | 0.80, 1.05 | 1.04 | 0.98, 1.21 |
| **Parents know someone who died of COVID (ref:No)** | 0.99 | 0.87, 1.14 | **1.13** | **1.01, 1.27** | **1.18** | **1.03, 1.36** |
| **Income loss due to childcare (ref:No)** | 0.87 | 0.69, 1.04 | 0.94 | 0.81, 1.08 | **0.79** | **0.65, 0.95** |
